# Possum faeces, the ‘canary in the coalmine’ heralding the emergence of Buruli ulcer in an Australian urban centre

**DOI:** 10.1101/2024.05.03.24306731

**Authors:** Bridgette J. McNamara, Jack Cornish, Kim R. Blasdell, Eugene Athan, Naomi E. Clarke, Tiffany Pe, Mohammad Akthar Hussain, Michael Muleme, Ee Laine Tay, Michael Dunn, Victoria Boyd, Anjana Karawita, Daniel P. O’Brien

## Abstract

Buruli ulcer (BU) endemicity is changing in Victoria, Australia. This study describes the emergence of BU cases in urban Geelong and examines timing and proximity of human cases to environmental *Mycobacterium ulcerans* DNA detection in possum faeces. From 2011-2022, 80 BU cases were reported, with clusters emerging in new endemic areas from 2019 onwards. Temporal analysis revealed cases occurred near *M. ulcerans* positive possum faeces, that preceded human cases by up to 39 months. Median diagnosis delay in these emerging areas was 7 weeks, longer than in established BU endemic areas. The findings emphasise the geographic and temporal association between BU emergence and possum faecal *M. ulcerans* presence, suggesting monitoring possum faeces as a viable early warning sign of increased BU risk in such areas. This proactive approach can enable early action to improve clinician and community BU awareness, or development of innovative control strategies, to reduce disease burden.

**Article Summary Line:** Our study describes the timing and pattern of Buruli Ulcer emergence in a non-coastal urban setting in Victoria, Australia, providing evidence that the emergence of clustered, locally acquired human cases was preceded by *Mycobacterium ulcerans* infection in possums up to three years prior.

**Author biography:** Dr. McNamara is an epidemiologist at the Barwon South West Public Health Unit and honorary senior research fellow at the Centre for Epidemiology and Biostatistics at the University of Melbourne. Her research interests focus on social and health equity and applied epidemiology for disease outbreak management and primary prevention, particularly for Buruli ulcer and COVID-19.

## INTRODUCTION

Buruli ulcer (BU) is a neglected tropical disease and remains an important public health issue across south-eastern Australia[1]. Manifesting as a necrotising ulcer, BU is caused by *Mycobacterium ulcerans* infection. Though often successfully treated with prolonged combination antibiotic therapy, it can result in significant morbidity[2]. Cases have been reported in several Australian states, with coastal areas on the Mornington and Bellarine Peninsulas in Victoria the predominant outbreak centres[3]. Whilst global case numbers are falling, the ongoing epidemic in Victoria appears to be worsening[4, 5].

BU is a notifiable condition in Victoria, mandating clinicians and pathology services to report diagnoses within 5 days. In 2023, Victoria saw a record 362 notified cases, surpassing previous peaks in 2018 (340 cases) and 2022 (338 cases) and exceeding the average yearly cases of the previous decade by over fivefold[6]. BU’s endemic regions in Victoria have shifted westward from initial notifications in East Gippsland in the 1940s, to Philip Island, and then the Mornington and Bellarine Peninsulas[3, 7]. Historically, cases have been concentrated in coastal areas; however, more recently cases have emerged within non-coastal suburbs of Melbourne and Geelong. The reasons for this emergence and the involved environmental sequence remains undetermined, impeding public health interventions to reduce disease spread.

*M. ulcerans* is an environmental pathogen[8]. Phylogenetic analysis suggests its introduction to new Victorian areas occurred centuries ago, with temporal analysis indicating a lag time from the bacterium arriving in an area to the emergence of human cases of 7-9 years [9]. Transmission involves complex interactions between humans, vectors, animals and the environment. Human acquisition routes may include mosquito bites[10], direct trauma or contact with contaminated substrates, particularly in limb areas with exposed skin [8, 11–13]. Unlike in Africa, Australia shows evidence of zoonotic transmission, notably from native ringtail (*Pseudocheirus peregrinus)* and brushtail (*Trichosurus vulpecula)* possums[14, 15]. Possum faeces at a residential property positive for *M. ulcerans* DNA correlated with higher likelihood of BU among residents[16, 17]. Surveillance in possums enhances human case prediction[17], yet the timing of infection in possums related to the emergence of human cases has not been described.

Prompt recognition of new at-risk areas for BU is crucial for public health management. Early detection of *M. ulcerans* in the environment may enable interventions to reduce disease emergence in humans. Improving community and clinician BU awareness in new at risk areas can improve prevention behaviors and delayed diagnosis, avoiding management challenges [11, 18, 19].

This study describes epidemiological and surveillance evidence in non-coastal urban Geelong, Victoria, identifying emerging BU endemic areas. We examine human BU cases, in terms of demographics, clinical presentation, and diagnosis timing, alongside *M. ulcerans* positive possum faeces, to inform future investigations into new BU transmission areas.

## METHODS

### Study design and setting

We conducted a descriptive epidemiological study, including a spatio-temporal clustering analysis of BU cases to identify new endemic areas in suburbs of Geelong, a city of approximately 250,000 people in southwest Victoria, Australia. Negligible risk ethics approval for the study was granted by the Barwon Health Human Research Ethic Committee (Project 23/10).

### Case definition and data sources

This study included laboratory-confirmed and probable cases of BU, as previously described [20]. We extracted data on all BU cases notified to the Department of Health (DH) Victoria between 1 January 2011 and 31 December 2022, from the Public Health Events Surveillance System (PHESS) for cases with a residential address within the Barwon South West (BSW) Public Health Unit catchment in southwest Victoria. Cases were geo-coded to longitude and latitude and to the corresponding Australian Bureau of Statistics ABS 2016 Mesh Block geographic statistical area[21]. Mesh blocks are the smallest unit of statistical geography in the ABS Australian Statistical Geography Standard (ASGS), in this instance denoting areas with 4-84 persons and 3-40 dwellings. These were then aggregated for analysis to broader geographic areas within the ASGS, namely Statistical Area 1 (SA1) and Statistical Area 2 (SA2) [21]. SA2 allow descriptions that equate loosely to known suburbs while having accurate GIS mapping boundaries and derivation of population estimates over time. The following central Geelong locations (SA2) and 2020 estimated residential populations were the focus of our study: Belmont (14,829 residents), Corio-Norlane (27,622), Geelong (13,781), Geelong West-Hamlyn Heights (21,272), Grovedale (31,579), Highton (23,869), Newcomb-Moolap (15,089), Newtown (10,945) and North Geelong-Bell Park (15,757) [22] (Supplementary Figure 1). Known proximal endemic areas for comparison included the following SA2: Ocean Grove -Barwon Heads, Point Lonsdale - Queenscliff, and Portarlington on the Bellarine Peninsula and Lorne-Anglesea on the Surf Coast (Figure 1).

**Figure 1.**
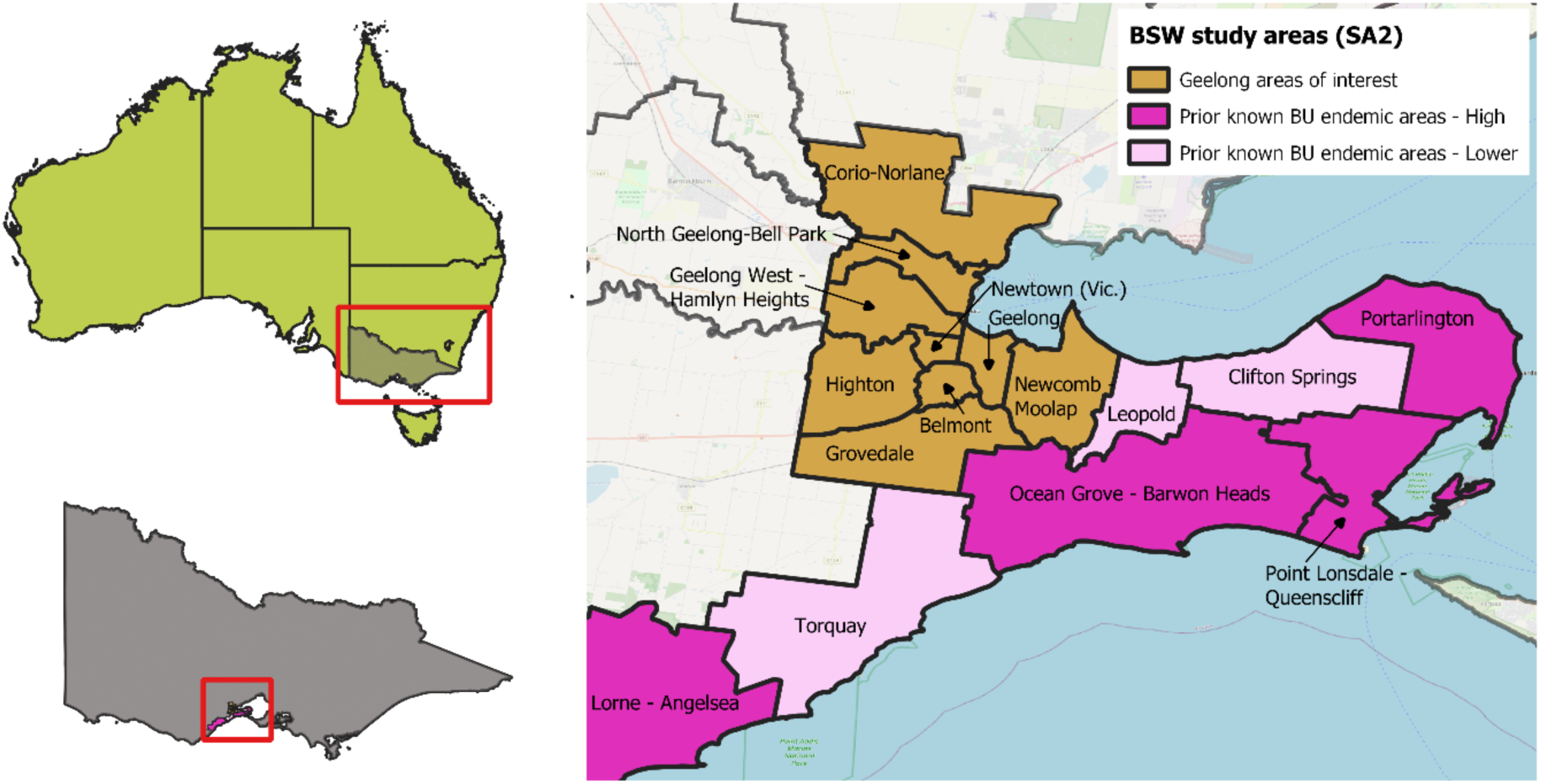
Location of central Geelong suburbs (Statistical Area 2 (SA2), brown) in Victoria, Australia, and prior known endemic areas in the Bellarine Peninsula and Surf Coast (higher case numbers dark pink, lower case numbers in light pink). Case numbers in dark pink (higher incidence areas) were >10 cases per 100,000 population in 2017-2019. Belmont with the central Geelong suburbs also meets this criterion.

### Epidemiology of cases

We used descriptive statistics to summarise age, sex, size of the BU lesion(s), season of symptom onset, time between symptom onset and diagnosis (based on case notification date), time between symptom onset and first healthcare presentation (*presentation delay*), and time between first healthcare presentation to diagnosis (*diagnosis delay*). For proportions, we calculated 95% confidence intervals using the binomial distribution. We examined reported travel to other endemic areas (as reported by treating clinicians and/or cases) and classified this to determine-those with either single or multiple visits to proximal (Bellarine or Surf Coast areas) or more distant known endemic regions of Victoria and those without. Differences in these characteristics across time periods (2011-2019, 2020-2022) and presentation and diagnosis delays were assessed using Chi-square tests or tests for difference of median values.

### Geographic distribution of BU cases over time

We examined aggregate case numbers and incidence per year at SA2 for the central Geelong suburbs and known endemic areas of the Bellarine and Surf Coast. Case numbers and case incidence at Statistical Area 1 (SA1) in central Geelong were mapped to visualise local clustering and the spread of cases, and changes in case incidence (total BU cases per 100,000 population) over time, across four time periods: 2011-2013, 2014-2016, 2017-2019, 2020-2022. For these analyses, the resident population within each SA1 and SA2 was sourced from ABS Estimated Resident Population (ERP) for the areas of interest from 2011 to 2020[22]. Population estimates for 2021 and 2022, which were not available for these geographic boundaries at the time of analysis, were derived from the average population growth estimated by the ERP of the previous five years.

### Spatio-temporal analysis of clustering of human BU cases

We examined geographic clustering of cases in central Geelong suburbs in a space-time analysis that examined clustering of cases mapped to Mesh blocks over time. We used Poisson models in SatScan^TM^ (trademark of Martin Kulldorff) [23, 24] to scan across the geographic locations over time to detect areas and time periods where the number of cases significantly exceeded the expected number of cases for the population to test the null hypothesis that cases are equally geographically distributed across the region for the period. For each significant cluster, we have reported the observed number of cases, the relative risk compared to the expected number under the null hypothesis and level of statistical significance. We conducted a subsequent, additional SatScan ^TM^ spatio-temporal analysis that allowed partial overlapping clusters (cluster centres were not within other clusters) and with the time-period restricted to single year intervals to further explore how the spread of the tightly clustered human cases within the Belmont area had emerged.

### Timing and proximity of BU cases to possum faecal samples positive for *M. ulcerans* DNA

We used data from a systematic possum faecal survey conducted in Geelong suburbs in 2020 [17], as well as follow-up surveys that were conducted in a smaller subset of Geelong regions at three, six and 24 months after the initial survey (Supplementary Figure 1). Details of survey collection methods have been published previously [17]. In brief, samples were collected across grid coordinates located 200m apart. Samples were collected from the nearest publicly accessible location to the grid coordinate (e.g., nature strips, footpaths). Field researchers also noted the coordinates and the date where no samples were found. Collected samples were homogenised, extracted, and analysed by IS2404 PCR to detect *M. ulcerans* DNA as described previously [25].

We calculated the proximity of human cases notified in 2020-2022 to possum faecal samples positive for *M. ulcerans* DNA. We have reported the median and interquartile range of the distance between the most proximal positive possum samples and residences of BU cases, as well as the number and proportion of cases within varying distance radius (200 m and 500 m). This analysis of proximity was undertaken for all cases residing in areas of the 2020 possum survey, for cases in the identified clusters, and for cases residing within areas with possum faecal testing in 2022. We also calculated the timing of case diagnosis in relation to the most proximal faecal detection in months.

## RESULTS

### BU case numbers and epidemiology

From 2011 to 2022, 402 BU cases were reported in the BSW catchment, including 80 cases within the SA2 locations of interest. Increasing case numbers and incidence were observed in Belmont from 2017-2022 and in Highton and Newtown areas from 2020-2022 (Table 1, Figure 2A, Supplementary Figure 2). Demographics, epidemiology, and clinical characteristics of cases within the Geelong areas of interest in the period 2011-2019 compared to those notified in 2020-2022 are presented in Table 2. The age of cases did not differ between the two time periods; however the proportion of female cases trended higher in the period 2020-2022 compared to 2011-2019 (p=0.052). Winter was the main season of symptom onset in both time periods. The proportion of cases without travel to other endemic areas was higher, but not statistically significant, in the period 2020-2022 (42%; 95% CI 32, 51) compared to 2011-2019 (29%; 95% CI 20, 37). Of those reporting travel, all cases reported visiting endemic areas on the Bellarine Peninsula or Surf Coast; additionally, 36% of these also reported travel to other endemic areas in Victoria. Time from symptom onset to diagnosis was similar in 2011-2019 and 2020-2022 (median 7-8 weeks, Table 2), but greater than that in other previously recognised endemic areas of the BSW region (Table S1 and Figure S3). The duration from symptoms to first seeking healthcare was similar in both the Geelong suburbs of interest and known endemic areas (median and IQR: 2.9 (0.9, 6.0) and 3.0 (1.0, 5.8) weeks, p=0.63), while the time from healthcare presentation to diagnosis differed; median and IQR of 2.6 (1.0, 5.9) weeks compared to 1.0 (0.6, 2.7) weeks in prior endemic areas, p=0.002 (Table S1, Figure S3).

**Figure 2A:**
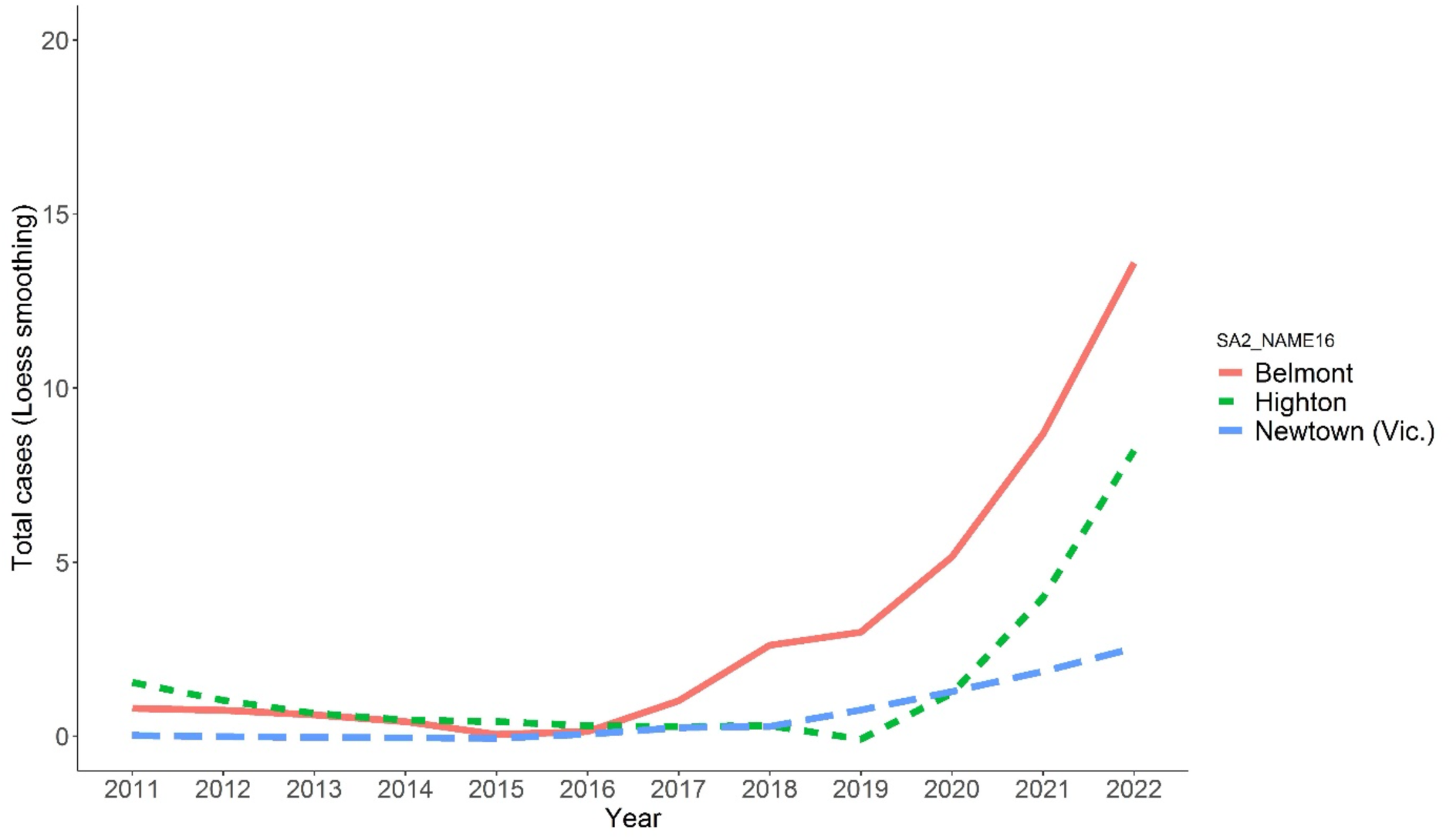
Trends in total cases numbers in SA2 with recent increases in transmission. Loess smoothing applied.

**Figure 2B:**
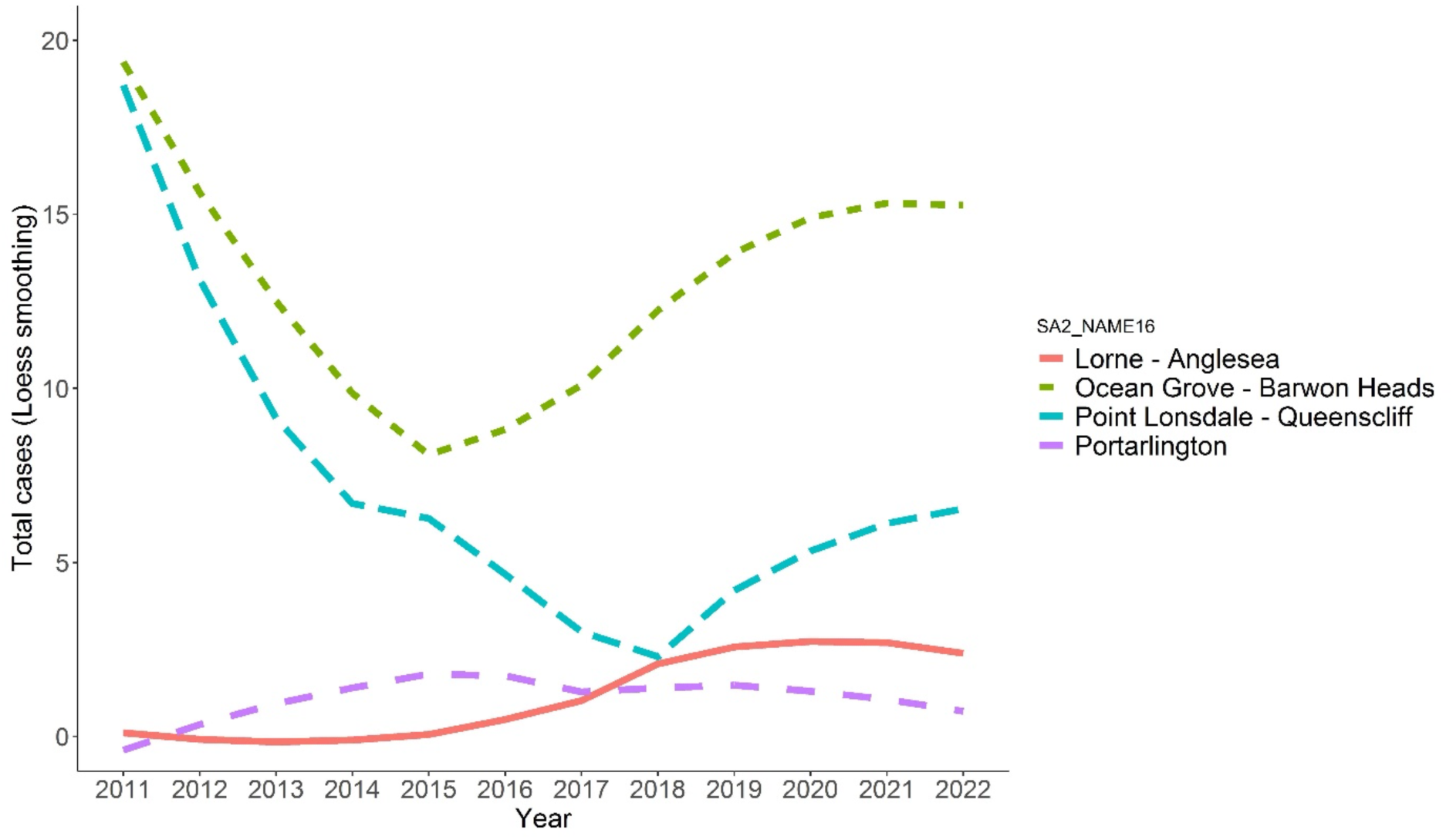
BU case numbers in SA2 previously recognised as endemic areas for BU with high case incidence. Loess smoothing applied.

**Table 1:** The number of cases and incidence in SA2 for Geelong regions of interest and SA2 of other recognised Bellarine endemic areas for the period from 2011-2017.

| SA2 | Measure | 2011-2013 | 2014-2016 | 2017-2019 | 2020-2022 |
| --- | --- | --- | --- | --- | --- |
| Belmont | Number of cases | 3 | 0 | 9 | 24 |
|  | Incidence (per 100,000py) | 7.2 | 0.0 | 20.5 | 53.5 |
| Corio - Norlane | Number of cases | 0 | 0 | 2 | 0 |
|  | Incidence (per 100,000py) | 0.0 | 0.0 | 2.4 | 0.0 |
| Geelong | Number of cases | 2 | 0 | 1 | 1 |
|  | Incidence (per 100,000py) | 5.3 | 0.0 | 2.5 | 2.4 |
| Geelong West - Hamlyn Heights | Number of cases | 2 | 0 | 0 | 2 |
|  | Incidence (per 100,000py) | 3.4 | 0.0 | 0.0 | 3.1 |
| Grovedale | Number of cases | 0 | 0 | 3 | 4 |
|  | Incidence (per 100,000py) | 0.0 | 0.0 | 3.6 | 4.0 |
| Highton | Number of cases | 3 | 1 | 1 | 12 |
|  | Incidence (per 100,000py) | 4.9 | 1.5 | 1.4 | 16.6 |
| Newcomb - Moolap | Number of cases | 3 | 0 | 0 | 0 |
|  | Incidence (per 100,000py) | 6.7 | 0.0 | 0.0 | 0.0 |
| Newtown (Vic.) | Number of cases | 0 | 0 | 1 | 6 |
|  | Incidence (per 100,000py) | 0.0 | 0.0 | 3.1 | 18.1 |
| North Geelong - Bell Park | Number of cases | 0 | 0 | 0 | 0 |
|  | Incidence (per 100,000py) | 0.0 | 0.0 | 0.0 | 0.0 |
| **Bellarine endemic areas** |  |  |  |  |  |
| Ocean Grove - Barwon Heads | Number of cases | 45 | 28 | 37 | 47 |
|  | Incidence (per 100,000py) | 76.8 | 41.0 | 46.2 | 51.0 |
| Point Lonsdale - Queenscliff | Number of cases | 38 | 20 | 7 | 21 |
|  | Incidence (per 100,000py) | 303.3 | 157.9 | 53.0 | 144.3 |
| Portarlington | Number of cases | 0 | 5 | 4 | 4 |
|  | Incidence (per 100,000py) | 0.0 | 22.8 | 16.8 | 15.2 |
| Lorne-Angelsea | Number of cases | 0 | 0 | 6 | 8 |
|  | Incidence (per 100,000py) | 0.0 | 0.0 | 37.1 | 46.8 |

The number of cases and incidence for the period in Geelong suburbs and comparisons with known endemic areas of the Bellarine (shaded grey). Incidence >10 per 100,000 person years (py) are shaded yellow. Aireys Inlet (7 cases since 2017); Anglesea (5 cases since 2017).

**Table 2:** Epidemiologic and clinical characteristics of BU cases in Geelong suburbs during each of the time periods and overall.

|  | 2011-2019 |  | 2020-2022 |  | Total |  | p-value |
| --- | --- | --- | --- | --- | --- | --- | --- |
|  | N=31<br>n | % (95%CI) | N=49<br>n | % (95%CI) | N=80<br>n | % (95%CI) |  |
| **Age (years)** |  |  |  |  |  |  | 0.95 |
| 0-19 | 4 | 13 (6, 19) | 6 | 12 (6, 19) | 10 | 13 (6, 19) |  |
| 20-39 | 9 | 29 (20, 38) | 12 | 24 (16, 33) | 21 | 26 (18, 35) |  |
| 40-59 | 9 | 29 (20, 38) | 14 | 29 (20, 37) | 23 | 29 (20, 38) |  |
| 60+ | 9 | 29 (20, 38) | 17 | 35 (25, 44) | 26 | 33 (23, 42) |  |
| Median (IQR) |  | 40 (29, 61) |  | 44 (31, 65) |  | 42 (29, 63) |  |
| **Sex** |  |  |  |  |  |  | 0.05 |
| Female | 10 | 32 (23, 41) | 28 | 57 (47, 67) | 38 | 48 (38, 57) |  |
| Male | 21 | 68 (59, 77) | 21 | 43 (33, 53) | 42 | 53 (43, 62) |  |
| Other | 0 |  | 0 |  | 0 |  |  |
| **Season of symptom onset\*** |  |  |  |  |  |  | 0.47 |
| Summer | 4 | 13 (6, 19) | 4 | 8 (3, 14) | 8 | 10 (4, 16) |  |
| Autumn | 8 | 26 (17, 34) | 7 | 14 (7, 21) | 15 | 19 (11, 26) |  |
| Winter | 13 | 42 (32, 52) | 27 | 55 (45, 65) | 40 | 50 (40, 60) |  |
| Spring | 6 | 19 (12, 27) | 11 | 22 (14, 31) | 17 | 21 (13, 29) |  |
| **Time from symptom onset\* to notification (weeks)** |  |  |  |  |  |  | 0.96 |
| Median (IQR) | 21 | 7.0 (5.7, 8.4) | 44 | 7.6 (4.1, 13.0) | 65 | 7.4 (4.9, 12.3) |  |
| **Lesion size: WHO classification** |  |  |  |  |  |  | 0.61 |
| Category 1 | 19 | 61 (52, 71) | 34 | 69 (60, 78) | 53 | 66 (57, 76) |  |
| Category 2 | 4 | 13 (6, 19) | 5 | 10 (4, 16) | 9 | 11 (5, 17) |  |
| Category 3 | 1 | 3 (0, 7) | 5 | 10 (4, 16) | 6 | 8 (2, 13) |  |
| Missing | 7 | 23 (14, 31) | 5 | 10 (4, 16) | 12 | 15 (8, 22) |  |
| **Travel to endemic regions outside area of residence** |  |  |  |  |  |  | 0.25 |
| No travel | 8 | 29 (20, 37) | 20 | 42 (32, 51) | 28 | 37 (27, 46) |  |
| Travel to endemic areas outside Geelong suburbs | 20 | 71 (63, 80) | 28 | 58 (49, 68) | 48 | 63 (54, 73) |  |
| Missing exposure data | 3 |  | 1 |  | 4 |  |  |

\*Missing first symptom onset (pre 2020, n=10; 2020-2022 n=5), these individuals were excluded from time from symptoms to notification, season of notification was used instead for season of symptom onset \*Chi-square on cases with data, missing excluded. World Health Organisation (WHO) lesion severity classification [37]: *Category 1* - single lesion <5cm diameter; *Category 2* - single lesion 5-15cm diameter; *Category 3* - a single lesion >15cm diameter, multiple lesions, lesions at a critical site (e.g. eye) or osteomyelitis.

### Distribution of human BU cases over time and proximity to possum faecal samples positive for *M. ulcerans* DNA

Spatio-temporal analyses identified three clusters suggesting local transmission in Geelong suburbs; Belmont (a cluster of 22 cases in 2019-2022), Highton (11 cases in 2022) and Newtown (3 cases in 2020-2021), with case numbers substantially higher than would be expected if cases followed a random geographic distribution (Figure 3). We observed southward and westward spread of human BU cases, with spread from an initial cluster in Belmont in 2019 (with up to five cases within a single SA1, Figure 4). The changing distribution of cases mirrored the changing distribution of positive possum faeces detected in the 2020 survey and the subsequent 2022 limited survey. Additional spatio-temporal analysis with overlapping clusters and single year periods confirmed the direction spread of BU cases (Supplementary Figure S4).

**Figure 3:**
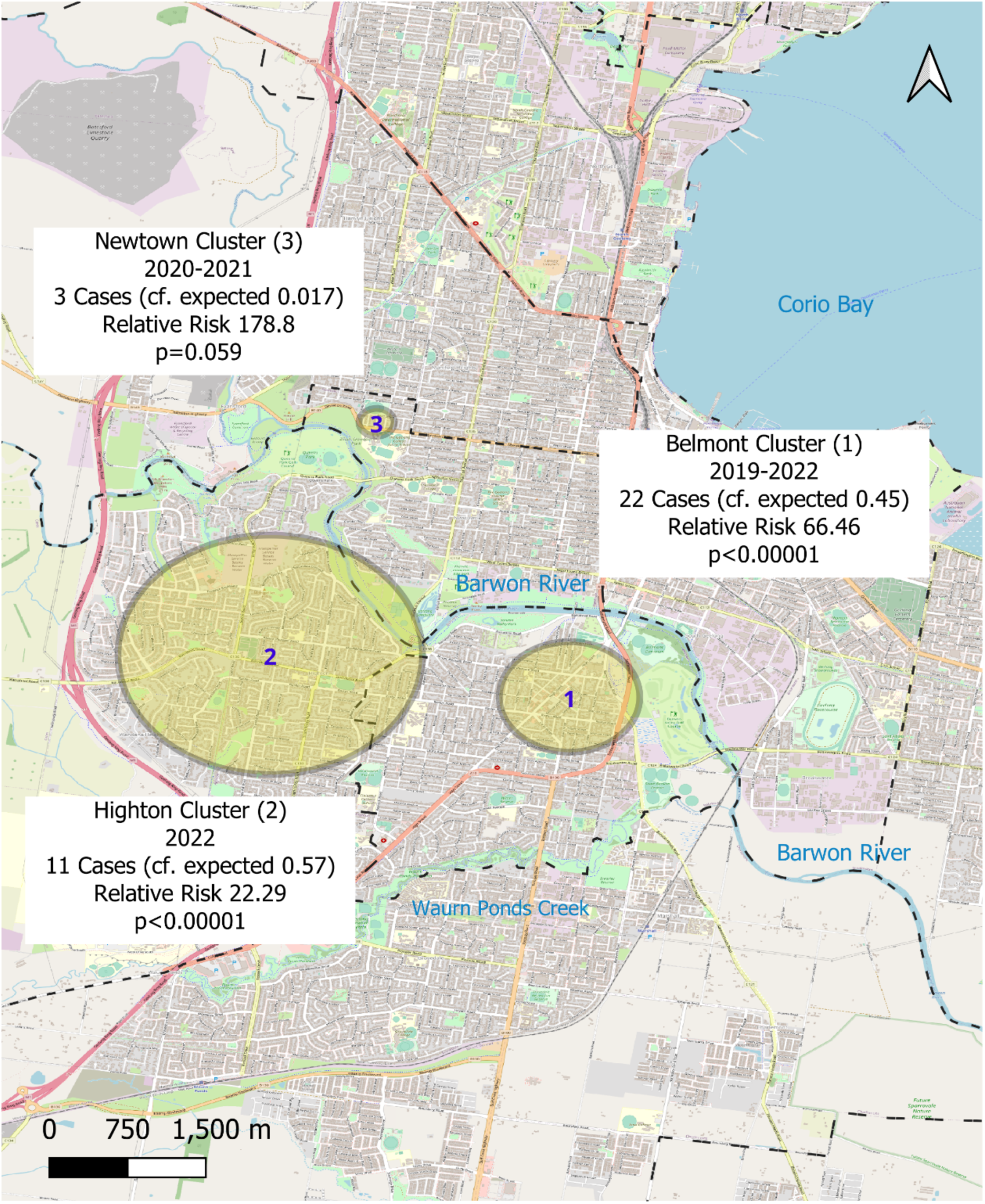
Map of urban Geelong clusters of BU cases in the period 2011-2022 identifed in spatio-temporal analysis (SatScan^TM^), in relation to the Barwon River (*Barra Warre N Yallok*), Waurn Ponds Creek (*Wuurrn Yallok*), Corio Bay (*Jillong*) and green space (parks and reserves).

**Figure 4:**
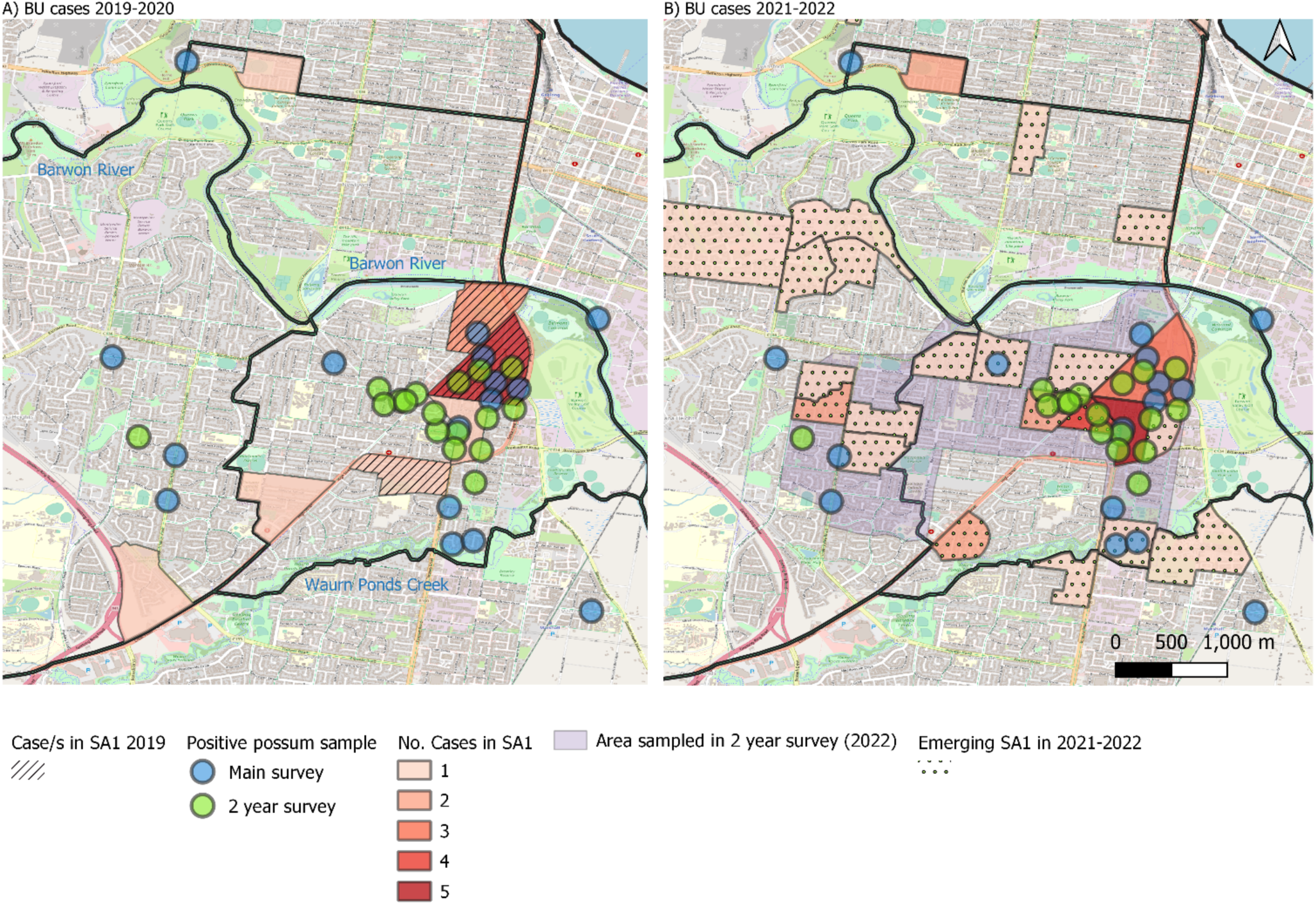
Distribution and timing of BU cases in 2019-2020 (A) and 2021-2022 (B) compared to the distribution of *M. ulcerans*-positive possum faecal samples collected in the main survey (2020) and the two-year follow up survey in 2022 (restricted area, indicated in light purple). Positive faecal samples from 2020 (blue) and 2022 (green) are presented with a 100m radius around the collection location. Case location of residence are visualised at Statistical Areas SA1, with the shading intensity indicating the number of cases during each period. SA1 in Figure 4A with cases in 2019 are highlighted with diagonal patterning, and areas with cases only in 2021-2022 are indicated with dotted pattern.

For the 39 BU cases diagnosed from 2020 and residing within areas included in the 2020 possum excreta survey the median distance from case residence to *M. ulcerans*-positive possum faeces was 283 metres. In all, 16/39 (41%) of the cases resided within 200 metres of a positive possum sample. The median distance to a positive possum sample was lower for cases within the identified clusters in Belmont, Highton and Newtown (199m; 50% of cases resided within 200 metres of a positive possum sample) than cases who were not within the identified clusters (median 1177m; 23% within 200m; p=0.040 rank-sum). For cases within the central areas sampled in both 2020 and 2022 faecal surveys, the median distance from a case residence to a prior positive possum sample was only 108m (IQR 82, 263).

The most proximal detections of *M. ulcerans* in possum faecal samples preceded the emergence of human BU cases by up to 39 months (IQR 8 -29months, Table 3). For cases in identified clusters, the most proximal positive possum faecal samples to cases occurred with an interquartile range of 7-31 months prior to the case diagnosis, apart from two cases where the closest positive detection (<100m) occurred after the case diagnosis. For these two cases, other positive possum faeces within 250m of the case residence were detected 16 and 18 months prior to the case diagnosis.

**Table 3:** Proximity and timing of human cases in surveyed areas with with *Mycobacterium ulcerans –* positive possum faecal samples (detection by IS2404 assay) collected in 2020 and 2022.

|  | Cases diagnosed<br>2020 onwards in<br>the main surveyed<br>areas | Cases within<br>detected<br>clusters* | Cases not in detected<br>clusters | Cases restricted to<br>those located in 2<br>year follow-up<br>survey area |
| --- | --- | --- | --- | --- |
| **Total number of cases** | 39 | 26 | 13 | 24 |
| **Distance from positive possum**<br>**faecal sample (m)** |  |  |  |  |
| Median (IQR) | 283 (92, 1025) | 199 (88, 393) | 1177 (209, 1256) | 108 (82, 263) |
| Range | 35, 1867 | 35, 1531 | 70, 1867 | 35, 405 |
| **Proximity** |  |  |  |  |
| <= 200m (n, %) | 16 (41%) | 13 (50%) | 3 (23%) | 16 (67%) |
| <=500m (n, %) | 26 (67%) | 20 (77%) | 6 (46%) | 24 (100%) |
| **Timing of positive possum**<br>**detections prior to cases** |  |  |  |  |
| median(IQR), months^ | 17 (8, 29) | 19 (8, 31) | 16 (11, 26) | 12 (7, 28) |

\*Clusters here include the two significant clusters (Belmont and Highton) and the Newtown cluster. All IS2404 positive faecal samples from any of the surveys (main 2020 survey, 3-month, 6-month or 24-month follow-up) were included in the measurement of proximity, with the closest sample contributing to these summary measures. ^month calculated as days divided by 30. Two cases with closest proximity samples <100m in 2022 (post diagnosis in 2021) have been excluded from this time calculation, although both had relatively proximal detections (within 250m) prior to diagnosis also.

Of note, we observed few BU cases in the northern suburbs of Geelong from 2011-2022 where possum population density was also lower evidenced by few to no possum faeces present during the 2020 excreta survey (Supplementary Figures S1 and S3); no cases were reported in North Geelong-Bell Park SA2, while the two cases reported in 2018 in Corio-Norlane SA2 resided within 344m and 1176m of the subsequent single *M. ulcerans*-positive possum faecal detection in 2020.

## DISCUSSION

BU endemicity in Victoria, Australia, is evolving. Our multidisciplinary investigation of human BU cases and *M. ulcerans* shedding in local possums, describes the speed and pattern of spread in a large urban, non-coastal centre in Victoria, Australia. Since 2017, BU cases have increased and clustered within three Geelong suburbs, predominantly near possums shedding *M. ulcerans* in faeces. Possum detection precedes human cases by months to years, suggesting that possum faecal surveillance for *M. ulcerans* may be useful in identifying emerging areas prior to onset of human cases. Finally, we also confirm delays to diagnosis in emerging BU areas compared to established endemic ones, highlighting the need for public health action and BU awareness.

Local transmission in Geelong is evident since 2017, despite the gaps in understanding BU transmission and the long incubation period complicating identification of where disease acquisition. Geelong residents frequently travel to the Bellarine Peninsula, especially in summer when the risk of *M. ulcerans* exposure peaks[26]. Enhanced surveillance by DH Victoria and the BSW Public Health Unit, shows that over 50% of cases in these areas had a travel history to known endemic areas during their exposure window. The high volume of such travel makes it difficult to completely discount its impact on our findings. However, a more random distribution of infections across Geelong would be expected if travel were the primary factor.

The tight clustering of suburban cases, particularly in locations where *M. ulcerans* was previously detected in possum excreta, suggests increased local acquisition. Since 2020, there has also been a slightly higher proportion of suburban cases without reported travel to other endemic areas compared with 2011-2019. Region-specific COVID-19 lockdowns may have influenced this difference in travel history, supporting more disease acquisition within Geelong during this time. Pandemic-related behavioural changes may have also affected the sex distribution of the Geelong cases, with a higher proportion of females in 2020-2022 (57%) compared to a higher number of males in 2011-2019 (69%), aligning with the known male predominance among BU cases in Victoria [27, 28].

While we provide clear evidence of emerging and increased transmission in several Geelong suburbs, the reasons for BU emergence in this region are unclear. Our data indicate that *M. ulcerans* likely entered Belmont, the first apparent outbreak centre before 2017. However, the manner of introduction and the environmental conditions sustaining bacterial prevalence for human transmission remains unknown.

Areas of Geelong with emerging human cases contain environmental factors previously shown to confer risk of BU transmission [16, 26]. The most important of these is that the environments of Belmont, Highton, and Newtown provide an ideal urban habitat for native possum species. Possum faeces positive for *M. ulcerans* have previously been correlated with nearby human cases [15, 16]. It appears clear that possums play a role in the BU transmission pathway in Victoria, as a mammalian reservoir of *M. ulcerans* bacteria [15, 25, 29]; possums manifest clinical disease and shed large quantities of viable *M. ulcerans* in their faeces [15, 25]. In prior Victorian outbreak areas, up to 40% of possum faeces tested positive for *M. ulcerans* DNA, with up to 24% of observed possums showing signs of clinical disease [15, 25]. Cases within the distinct urban clusters in our study all occurred near positive possum faecal samples collected approximately 8-31 months prior to the case diagnosis.

Importantly, few human cases were identified in areas without positive possum faecal samples, despite some limitation to the surveying areas. This aligns with prior findings of lower BU risk in areas without positive faecal samples [16]. In addition, clusters of cases have not emerged in the northern suburbs of Geelong where a relative absence of possums in these areas was suggested by an inability to find possum faecal specimens in the main survey of 2020 (Supplementary Figure 1).

Further, other environmental conditions in these areas may be contributing to increased transmission; streets within these Geelong suburbs are well lined with large trees and overhead power lines that facilitate possum mobility[25] (Supplementary Figure 5), and many are in proximity to parkland areas. Additionally, the larger Belmont cluster is adjacent to a major local waterway, the Barwon River (*Barra Warre N Yallok*), as well as low-lying areas prone to flooding. Of note, flooding occurred in Highton in January 2022, coinciding with the peak exposure period for infection. The influence of rainfall on BU transmission has been identified previously, with an approximate lag time of 10-12 months from significant rainfall events to an increase in cases[26]. It is hypothesised the majority of disease acquisition occurs in warmer months, as a result of optimal climatic conditions, changes in human behaviour, and increased abundance of potential vector species (mosquitoes), and higher likelihood of inoculating events[10]. The majority of cases we analysed reported initial symptoms in winter and spring, aligning with known lag times and previous spring rainfall being well above average [30].

At this stage there are no proven public health interventions to limit the emergence of BU cases in Victoria, as evidenced by the continuing geographical spread of the disease. Additionally, information has been lacking relating to the environmental sequence of events involved in the emergence of the BU disease in previously non-endemic areas. Our research findings support the hypothesis that the disease emerges in possum populations prior to human populations. We have demonstrated *M. ulcerans* presence in the possum populations more than 2.5 years prior to the emergence of clustered human cases with the spread of BU cases following the direction of positive detections in possum faeces. Prior evidence from residential environmental surveys in the Mornington Peninsula, Victoria, and the current longitudinal data from Geelong suggests that *M. ulcerans* shedding in possum populations remains for extended periods at specific locations but can vary over time[25]. Our findings lend support to a recent publication of a statistical model to predict the location of future human BU cases based upon the location of positive possum faecal specimens [17]. However, in our study we extend the timeframe for case follow-up to quantify the time between the positive detections in possum populations and subsequent human cases, with indication that cases are likely within 3 years of the environmental detection. As such, future surveillance models using possum faecal positivity may provide an early warning signal (“canary in the coalmine”) to emerging areas at risk of human BU cases, as well as providing a tool to help delineate new areas of local transmission from potential travel-related acquisition in areas with current cases. Furthermore, the significant time delay between infections in possum populations and the emergence of human cases may allow the development of interventions that prevent BU disease occurring in residents or visitors to that area (e.g. local vaccination of possum populations[31] and/or significantly increased vector control[10]). However, further investigation into the practicalities around collection distances, intervals between collection surveys and overall cost-effectiveness is required. Current research into lower-cost field-based methods for detecting the bacterial DNA offers hope for increased feasibility of ongoing systematic faecal surveillance[32] that could better track the movement of the bacteria around the state prior to outbreaks in humans.

Managing BU in urban areas requires a One Health approach, incorporating environmental, wildlife and human factors. With possum populations well established in suburban Geelong, avoiding contact with contaminated environments is challenging. Culling possums is impractical given their protected status in Victoria, and past removal of reservoir hosts elsewhere resulted in paradoxical spread of disease [33]. New research focusing on infection control in possum populations, particularly vaccination, is promising[31]. Moreover, there is evidence supporting various basic protective measures to reduce risk of BU in humans[11, 34]. In a recent comprehensive case-control study, we demonstrated that residents of Victorian endemic areas who reported multiple skin protection and hygiene behaviours experienced 60%-80% lower odds of BU. These measures included wearing protective clothing outdoors, use of insect repellent in warmer months and immediate washing and/or applying antiseptic to any wounds obtained outdoors[11].

Rapid recognition of new BU transmission areas is vital for targeted prevention messaging and education to increase community and clinician awareness. Our study highlights delayed diagnosis post health care presentation in emerging transmission areas compared to established endemic regions. Diagnosis times varied widely, limited by missing data. However, delays in presentation and diagnosis are common in non-endemic areas[18], impacting treatment and clinical outcomes [35]. Surveillance of *M. ulcerans* in possum populations can facilitate early education of medical practitioners to improving diagnosis and treatment timeliness. Local Public Health Units, established in late 2020 in Victoria[36], are well positioned for community engagement to enhance awareness and clinician knowledge, reducing overall burden of disease.

## Data Availability

Sharing of de-identified human case data used in this publication may be requested from the Department of Health Victoria, contingent on approval of an analysis plan by this Department and relevant ethical approval. Possum faecal data may be shared, after publication, on a collaborative basis upon reasonable request made to Dr Kim Blasdell.

## Acknowledgements

The authors would like to acknowledge the work of the Public Health Officers (K. Cham, B. Eedara, S. Fletcher, J. FitzGibbon, S. Howard, L. Nolte, T. Pe, S. Thompson, L. Veenhuizen, R. Ward, I. Caple) and Team Leaders (L. Farnsworth, S. Hallissey, A. Kelly, J. O’Neill) within the BSW Public Health Unit for their follow-up of notified cases since July 2022 and assistance with the current epidemiological investigation, as well as the Department of Health Victoria, Health Protection team for follow-up of cases prior to 2022 and guidance in the response to the emerging areas. We acknowledge the field researchers at CSIRO involved in collection and testing of the possum faecal samples in Geelong. The data collection and analysis of the possum faecal surveys were funded as part of the NHMRC Partnership Project Grant 2018-2020 (GNT1152807) led by Professor Tim Stinear. Ethical approval for the current analyses were granted by the Barwon Health Human Research Ethics Committee under a negligible risk application (Project 23/10).

## Author contributions

The authors confirm contribution to the paper as follows: study conception and design: BM, JC, KB, DO; data collection: BM, TP, NC, ET (human); KB, MD, VB, AK (possum); analysis and interpretation of results: BM (epi analysis) VB, KB (faecal PCR analysis), JC, KB, EA, NC, TP, MAH, MM, DO (interpretation) ; draft manuscript preparation: BM, JC. All authors reviewed the results and approved the final version of the manuscript.

## Supplementary information

**Supplementary Figure S1:**
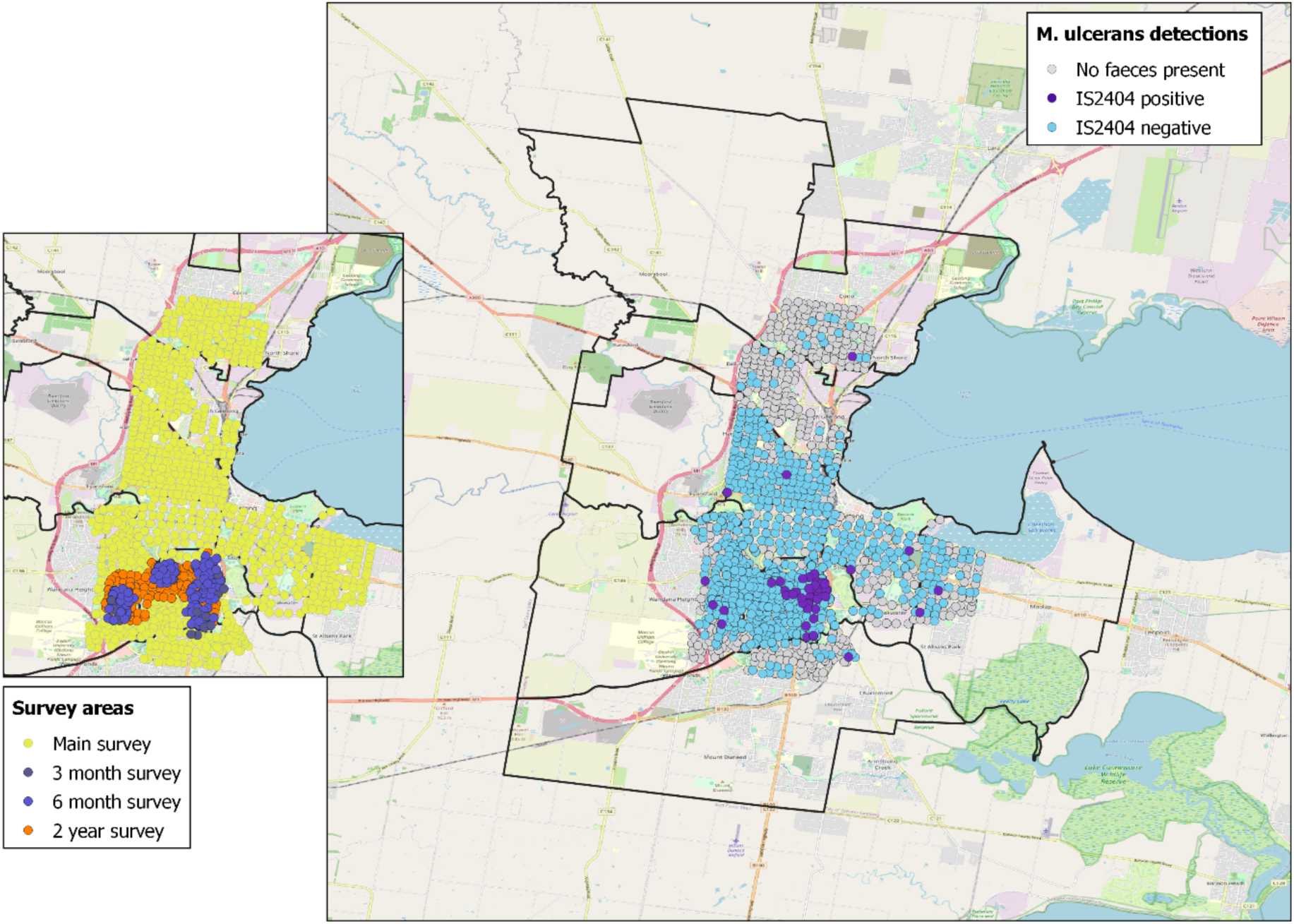
Map of possum faecal sample collection (inset) – the main survey (conducted Jan-April 2020, yellow; 3 month follow up (May-Jun 2020) and 6 month follow up (August –September 2020), purple; areas surveyed in the 24 month follow up (February to April 2022) in orange. Note 24 month survey areas overlay Main survey areas and 3- and 6-month areas. Map of possum faeces locations and *M. ulcerans* detections via IS2404 PCR (main map) – grey points were surveyed but no faeces were present.

**Supplementary Figure S2:**
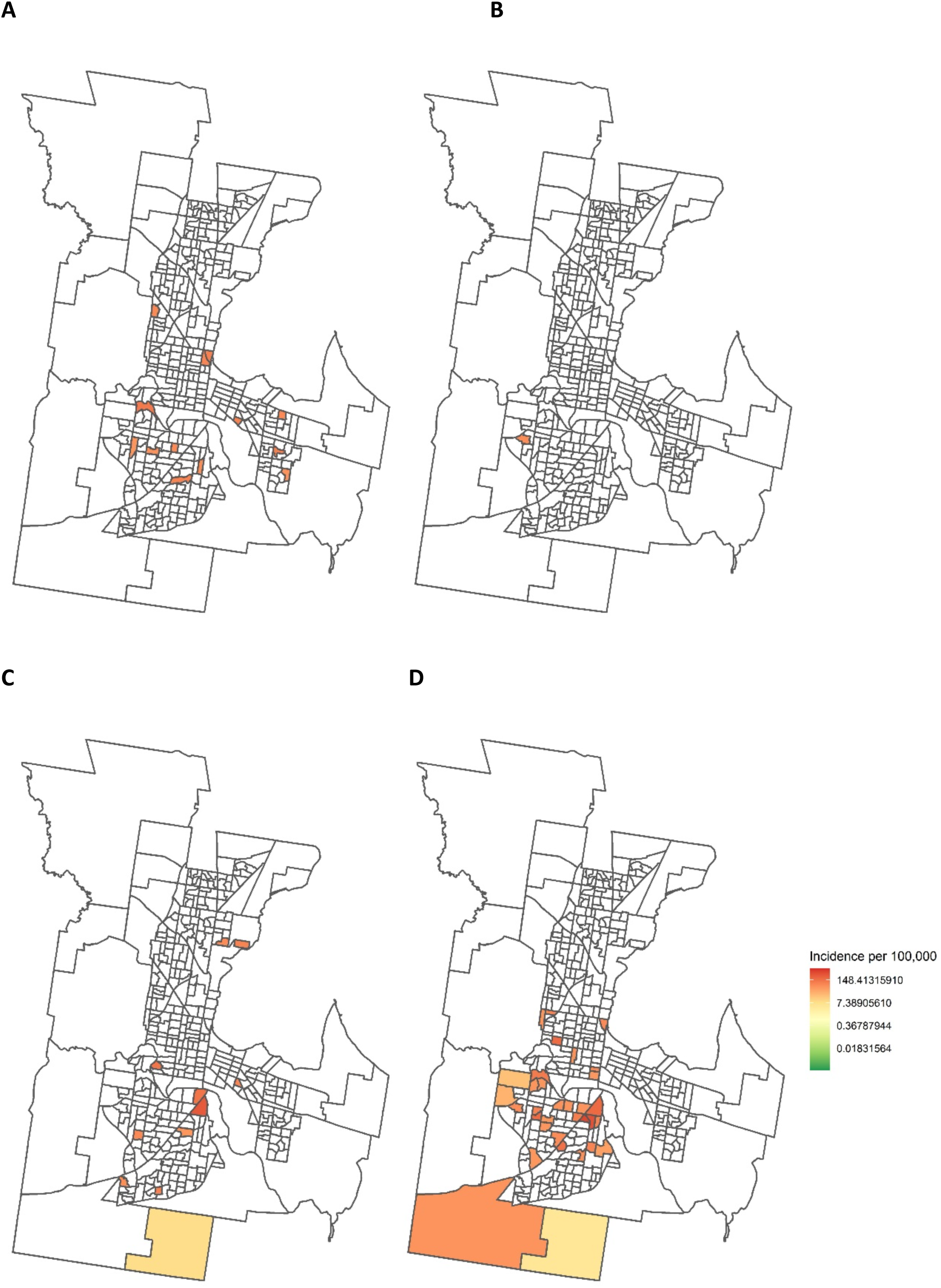
Geographic distribution of cases per 100,000 person-years (shown on log-scale) across the four time periods (A) 2011-2013, (B) 2014-2016, (C) 2017-2019 and (D) 2020-2022. Note areas with 0 cases shown in white; this also includes n=16 areas without any estimated residential population (i.e. parkland).

**Supplementary Figure S3:**
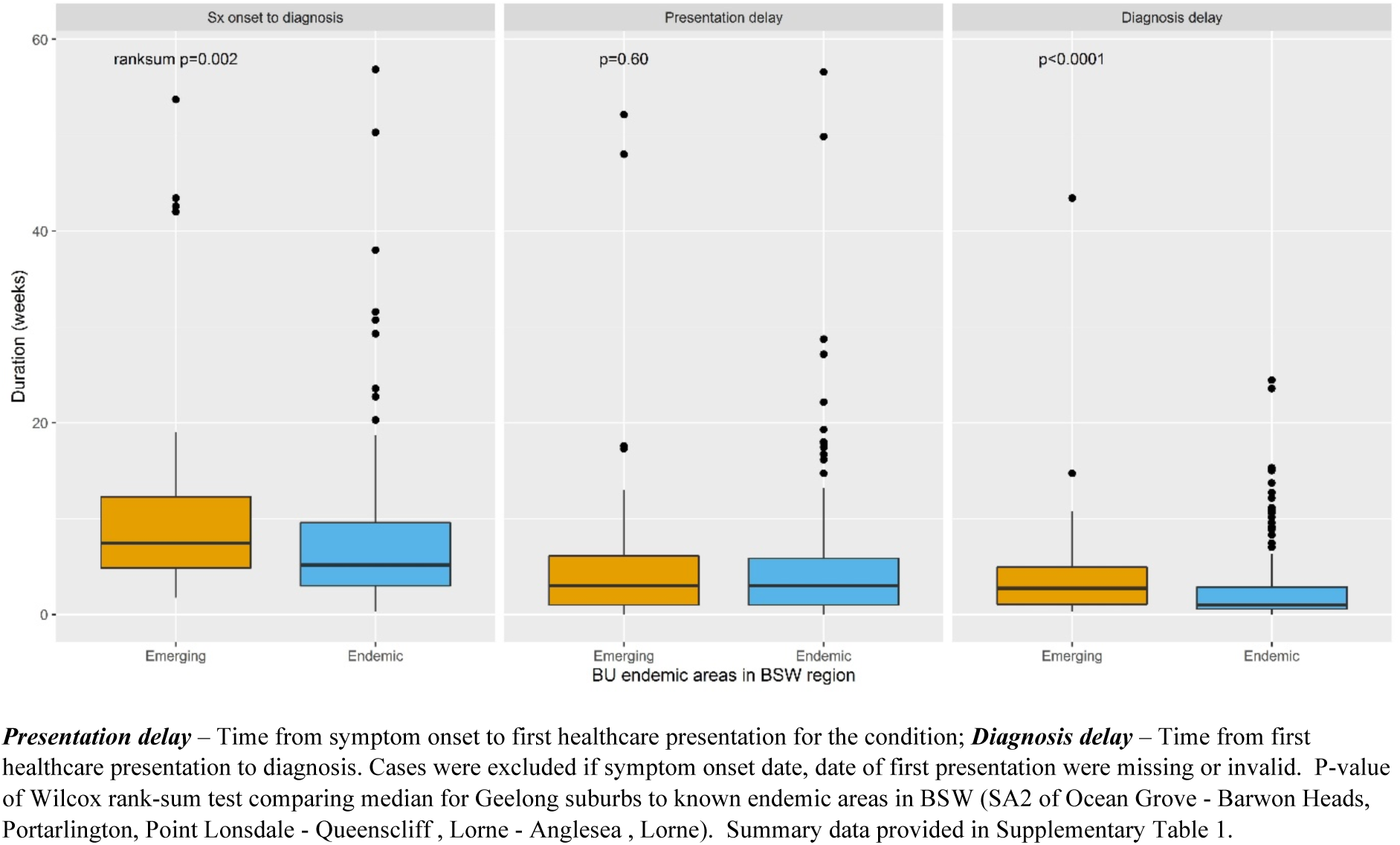
Time to BU diagnosis after symptom onset and presentation and diagnosis delays for BU cases in central Geelong suburbs from 2011-2022 compared to BU cases in previously known endemic areas of high case incidence in the Barwon South West Region of Victoria, Australia.

**Supplementary Table S1:** Timing BU diagnosis after symptom onset, and the presentation delay (time from symptom onset to first healthcare presentation) and diagnosis delay (time from healthcare presentation to diagnosis) for BU Cases in central Geelong suburbs notified 2011-2022 compared to BU cases in previously known endemic areas of the Barwon South West (BSW) Region of Victoria, Australia.

|  | *A) Geelong suburbs of interest (new areas)* |  | *B) Known endemic areas in BSW\** |  | **p-value**<br>**(A vs.B)<sup>#</sup>** |
| --- | --- | --- | --- | --- | --- |
|  | **N cases** | **Weeks**<br>**(median, IQR)** | **N cases** | **Weeks (median,**<br>**IQR)** |  |
| **Overall 2011-2022** |  |  |  |  |  |
| Time from symptoms to diagnosis | 65 | 7.4 (4.9, 12.3) | 189 | 5.1 (3.0, 9.6) | 0.002 |
| Presentation delay | 60 | 3.0 (1.0, 6.1) | 181 | 3.0 (1.0, 5.9) | 0.60 |
| Diagnosis delay | 59 | 2.7 (1.1, 4.9) | 195 | 0.9 (0.6, 2.9) | <0.0001 |
| **2011-2019** |  |  |  |  |  |
| Time from symptoms to diagnosis | 21 | 7.0 (5.7, 8.4) | 133 | 5.1 (3.0, 9.3) | 0.03 |
| Presentation delay | 18 | 2.9 (0.9, 6.0) | 127 | 3.0 (1.0, 5.8) | 0.63 |
| Diagnosis delay | 21 | 2.9 (0.9, 4.4) | 138 | 1.0 (0.6, 2.9) | 0.03 |
| **2020-2022** |  |  |  |  |  |
| Time from symptoms to diagnosis | 44 | 7.6 (4.1, 13.0) | 56 | 4.9 (3.4, 9.7) | 0.06 |
| Presentation delay | 42 | 3.0 (1.0, 6.3) | 54 | 2.3 (1.0, 6.0) | 0.66 |
| Diagnosis delay | 38 | 2.6 (1.1, 5.9) | 57 | 1.0 (0.6, 2.7) | 0.002 |

IQR: Interquartile Range. Cases were excluded if symptom onset date, date of first presentation were missing or invalid. P-value of Wilcoxon rank-sum test comparing Geelong suburbs to known endemic areas. \*Known endemic areas included SA2 of "Ocean Grove - Barwon Heads", "Portarlington", "Point Lonsdale - Queenscliff", "Lorne - Anglesea", "Lorne".

**Supplementary Figure S4:**
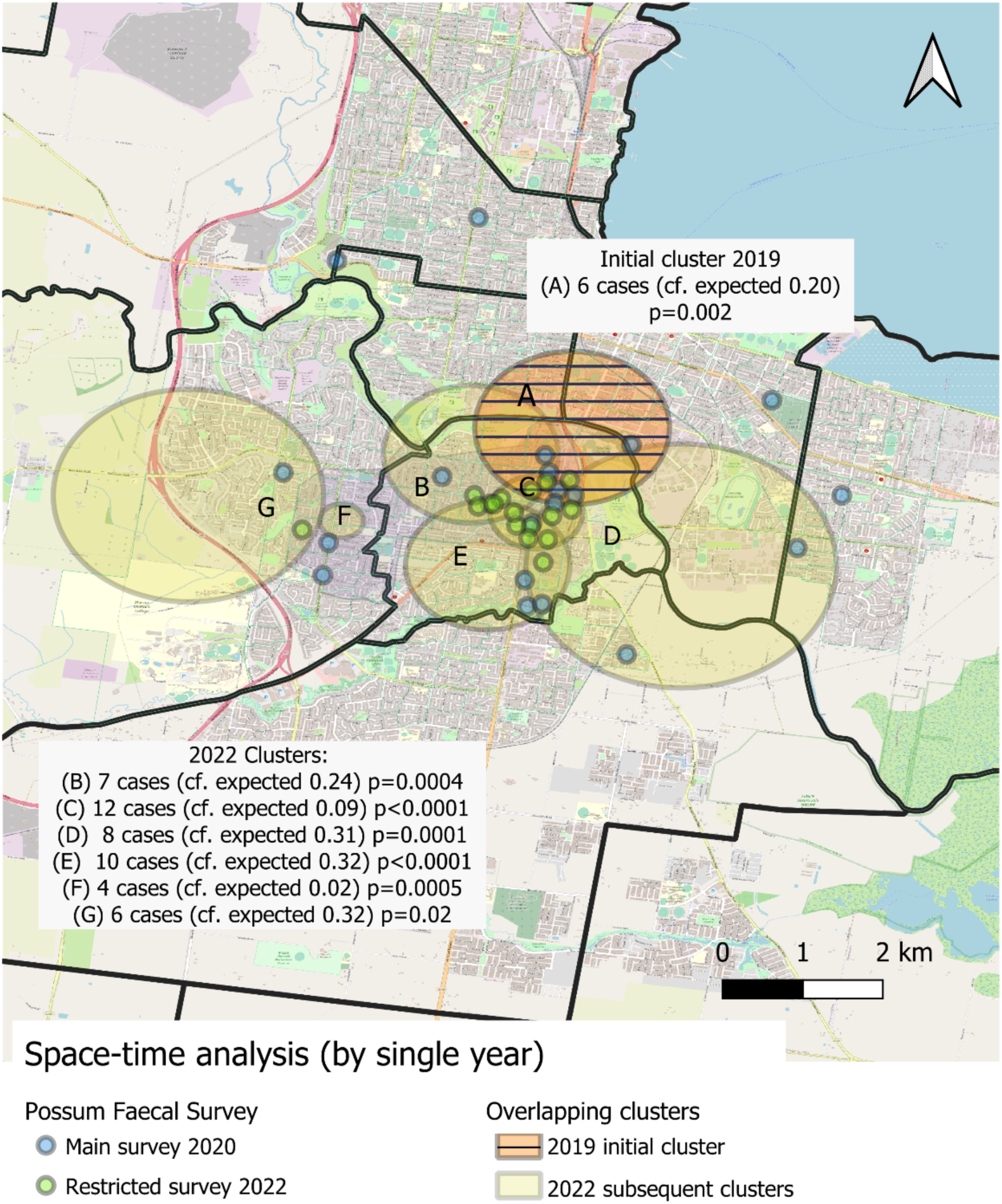
Spatio-temporal (SatScan^TM^) analysis (with overlapping case clusters and single year periods). Outward spread was observed from a central 2019 cluster (Cluster A, orange with striped pattern), with significant surrounding subsequent clusters identified in 2022 (Clusters B-G). The spread and increasing density of *M. ulcerans* in the native possum population is indicated by *M. ulcerans*-positive possum faecal samples from the main 2020 survey (blue) and the restricted 2 year follow up survey (green).

**Supplementary Figure S5:**
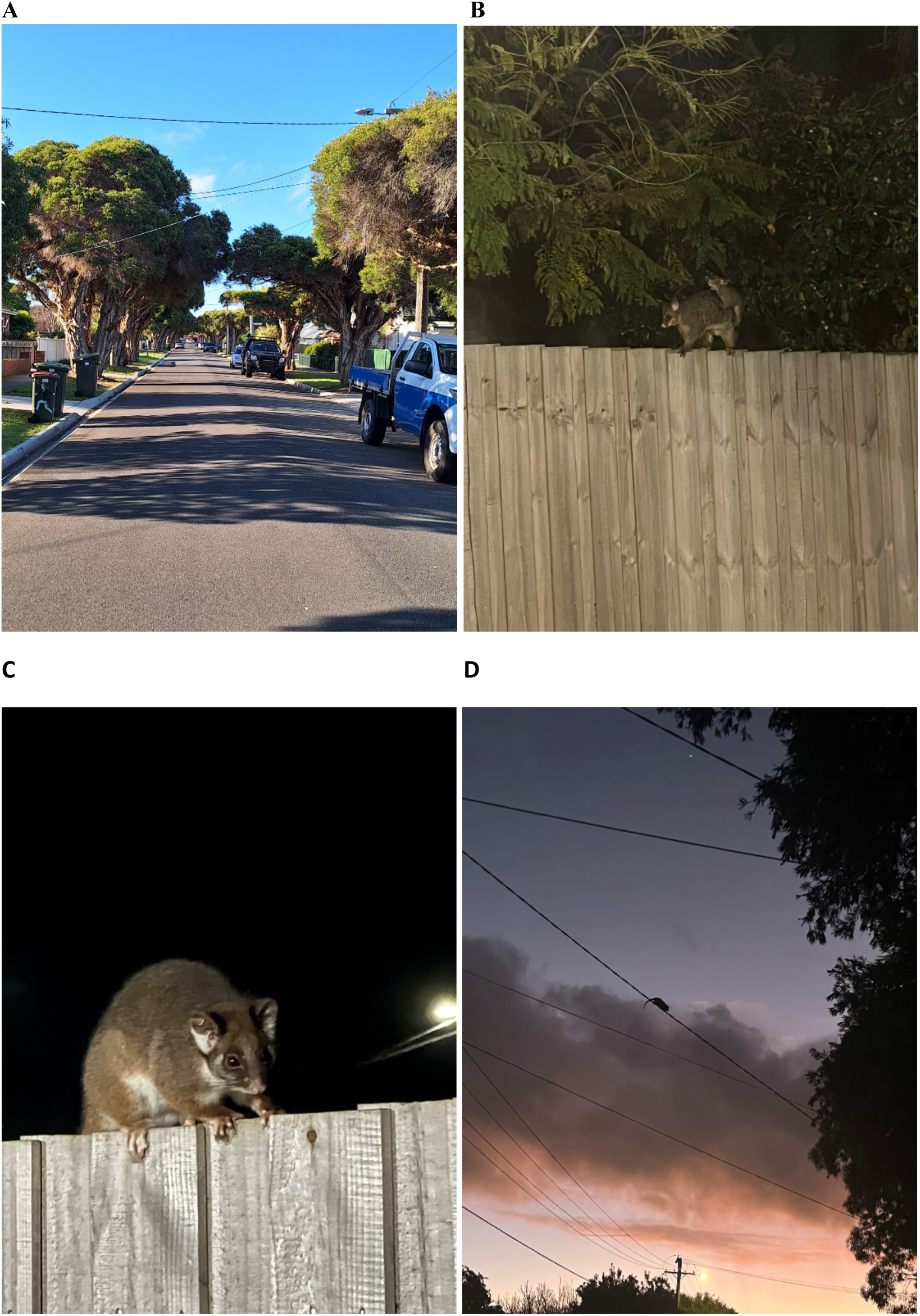
A) Photograph of typical Belmont street in Geelong showing trees and powerlines that aid in mobility of the local native possum population. B-D) Example photographs of the highly mobile possum population in Highton, City of Geelong.

## REFERENCES

1. World Health Organization, Working to Overcome the Global Impact of Neglected Tropical Diseases: First WHO Report on Neglected Tropical Diseases. 2010: Geneva, Switzerland.

2. Loftus, M.J., et al., A severe case of Mycobacterium ulcerans (Buruli ulcer) osteomyelitis requiring a below-knee amputation. Med J Aust, 2018. 208(7): p. 290–291.

3. Loftus, M.J., et al., *Epidemiology of Buruli Ulcer Infections, Victoria, Australia*, *2011-2016*. Emerg Infect Dis, 2018. 24(11): p. 1988–1997.

4. World Health Organization. Country data for Buruli ulcer. 2023 [cited 30th January 2023]; Available from: http://apps.who.int/neglected_diseases/ntddata/buruli/buruli.html.

5. O’Brien, D.P., et al., The changing epidemiology worldwide of Mycobacterium ulcerans. Epidemiol Infect, 2018. 147: p. e19.

6. Department of Health Victoria, Victoria, local public health areas and local government areas surveillance summary report *(accessed 4 Dec 2023)* https://www.health.vic.gov.au/infectious-diseases/local-government-areas-surveillance-report. 2023, Department of Health Victoria: Melbourne.

7. Mac, C.P., J.C. Tolhurst, and, et al., A new mycobacterial infection in man. J Pathol Bacteriol, 1948. 60(1): p. 93–122.

8. Merritt, R.W., et al., Ecology and transmission of Buruli ulcer disease: a systematic review. PLoS Negl Trop Dis, 2010. 4(12): p. e911.

9. Buultjens, A.H., et al., Comparative Genomics Shows That Mycobacterium ulcerans Migration and Expansion Preceded the Rise of Buruli Ulcer in Southeastern Australia. Appl Environ Microbiol, 2018. 84(8).

10. Mee, P.T., et al., Mosquitoes provide a transmission route between possums and humans for Buruli ulcer in southeastern Australia. Nat Microbiol, 2024.

11. McNamara, B.J., et al., Comprehensive Case–Control Study of Protective and Risk Factors for Buruli Ulcer, Southeastern Australia. Emerg Infect Dis, 2023. 29(10): p. 2032–2043.

12. Yerramilli, A., et al., The location of Australian Buruli ulcer lesions-Implications for unravelling disease transmission. PLoS Negl Trop Dis, 2017. 11(8): p. e0005800.

13. Lavender, C.J., et al., Risk of Buruli ulcer and detection of Mycobacterium ulcerans in mosquitoes in southeastern Australia. PLoS Negl Trop Dis, 2011. 5(9): p. e1305.

14. Röltgen K, P.G., Epidemiology and disease burden of Buruli ulcer: a review. Res Rep Trop Med, 2015. 6: p. 59–73.

15. Fyfe, J.A., et al., A major role for mammals in the ecology of Mycobacterium ulcerans. PLoS Negl Trop Dis, 2010. 4(8): p. e791.

16. Blasdell, K.R., et al., Environmental risk factors associated with the presence of Mycobacterium ulcerans in Victoria, Australia. PLOS ONE, 2022. 17(9): p. e0274627.

17. Vandelannoote, K., et al., Statistical modeling based on structured surveys of Australian native possum excreta harboring Mycobacterium ulcerans predicts Buruli ulcer occurrence in humans. Elife, 2023. 12.

18. Coutts, S.P., et al., *Delays in Patient Presentation and Diagnosis for Buruli Ulcer (Mycobacterium ulcerans Infection) in Victoria, Australia*, *2011-2017*. Trop Med Infect Dis, 2019. 4(3).

19. Tai, A.Y.C., et al., Increased Severity and Spread of Mycobacterium ulcerans, Southeastern Australia. Emerg Infect Dis, 2018. 24(1): p. 58–64.

20. Betts, J.M., et al., Buruli ulcer: a new case definition for Victoria. Commun Dis Intell (2018), 2020. 44.

21. ABS, 1270.0.55.001 - Australian Statistical Geography Standard (ASGS): Volume 1 - Main Structure and Greater Capital City Statistical Areas, July 2016 2017, Australian Bureau of Statistics: Canberra.

22. ABS, Regional Population, Estimated Residential Population 2001-2020. 2022, Australian Bureau of Statistics: Canberra.

23. Kulldorff, M., A Spatial Scan Statistic. Commun. Statist. - Theory Meth., 1997. 26(6): p. 1481–1496.

24. Kulldorff, M. and Information Management Services Inc., SaTScanTM v8.0: Software for the spatial and space-time scan statistics. http://www.satscan.org/, . 2009.

25. Blasdell, K.R., et al., Environmental risk factors associated with the presence of Mycobacterium ulcerans in Victoria, Australia. PLoS One, 2022. 17(9): p. e0274627.

26. Yerramilli, A., et al., The association of rainfall and Buruli ulcer in southeastern Australia. PLoS Negl Trop Dis, 2018. 12(9): p. e0006757.

27. McNamara, B., et al., Comprehensive case-control study of protective and risk factors for Buruli ulcer in South-Eastern Australia. Emerg Infect Dis, 2023. **(under review)**.

28. O’Brien, D.P., et al., Mycobacterium ulcerans in the Elderly: More Severe Disease and Suboptimal Outcomes. PLoS Negl Trop Dis, 2015. 9(12): p. e0004253.

29. Singh, A., et al., Potential Animal Reservoir of Mycobacterium ulcerans: A Systematic Review. Trop Med Infect Dis, 2018. 3(2).

30. BureauofMeteorology. *Climate Data Online*. 2022 [cited 2022 December].

31. O’Brien, D.P., et al., Is BCG vaccination of possums the solution to the Buruli ulcer epidemic in south-eastern Australia? Med J Aust, 2023. 219(11): p. 520–522.

32. World Health Organization, Ending the neglect to attain the Sustainable Development Goals: a strategic framework for integrated control and management of skin-related neglected tropical diseases. . 2022, World Health Organization: Geneva.

33. Donnelly, C.A., et al., Positive and negative effects of widespread badger culling on tuberculosis in cattle. Nature, 2006. 439(7078): p. 843–6.

34. Quek, T.Y., et al., Risk factors for Mycobacterium ulcerans infection, southeastern Australia. Emerg Infect Dis, 2007. 13(11): p. 1661–6.

35. Quek, T.Y., et al., Mycobacterium ulcerans infection: factors influencing diagnostic delay. Med J Aust, 2007. 187(10): p. 561–3.

36. Department of Health Victoria. *Local Public Health Units*. [Webpage] 2023 [cited 2024 5 Feb]; Available from: https://www.health.vic.gov.au/local-public-health-units.

37. World Health Organization, *Treatment of Mycobacterium Ulcerans Disease (Buruli Ulcer): Guidance for Health Workers*. . 2012, WHO Press: Geneva, Switzerland.

